# Spatial Optimization to Improve COVID-19 Vaccine Allocation

**DOI:** 10.1101/2022.10.30.22281737

**Authors:** Justin Goodson, Stephen Scroggins, Tasnova Afroze, Enbal Shacham

## Abstract

Early distribution of COVID-19 vaccines was largely driven by population size and did not account for COVID-19 prevalence nor location characteristics. In this study, we applied an optimization framework to identify distribution strategies that would have lowered COVID-19 related morbidity and mortality. Optimized vaccine allocation would have decreased case incidence by 8% with 5,926 fewer COVID-19 cases, 106 fewer deaths, and 4.5 million dollars in healthcare cost saved during the first half of 2021. As COVID-19 variants continue to be identified, and the likelihood of future pandemics remains high, application of resource optimization should be a priority for policy makers.

## Introduction

The COVID-19 pandemic has had a devastating effect worldwide. In the U.S. alone, by the end of 2020 there were more than 20 million reported infections, greater than 1.1 million related hospitalizations, and nearly 364 thousand related deaths.^1^ As demand for beds, medical personnel, and equipment quickly outpaced supply, hospitals turned away the ill and suspended preventive and elective procedures.^2,3^ While multiple COVID-19 vaccinations were developed at unprecedented speeds and made available to general adult U.S. populations by early 2021^4^, challenges in the distribution of limited vaccine supplies quickly arose.^5,6^

In the U.S., states received the bulk of vaccine supplies from the federal government in an amount typically proportional to their population size.^7^ U.S. states were then tasked with downstream distribution to residents and local agencies. Although consumer vaccine distribution in most states was preliminarily undertaken in phases, based on infection susceptibility and likelihood^8^, this, and subsequent distribution to the general adult population, was largely based on geographic population size. Though a local distribution method based on population size appealed to a sense of equity, it negated a typically more accepted needs-based approach. Currently, the possibility of optimal vaccine allocation during the early stages of a pandemic is not well understood.

Integrating classical models of infectious disease transmission into a vaccine allocation optimization framework is challenging. In typical compartmental models, the equations governing transitions among classes are non-convex. Incorporating these transitions into an optimization model results in complex and computationally challenging problems. In most cases, the allocations returned by state-of-the-art optimization techniques carry no guarantee of optimality.^9^ Further, the complexity of such methods restricts their accessibility and use in practice, an important requirement as COVID-19 continues to spread, causing morbidity and mortality. Towards that end, optimal vaccine allocation requires an understanding of population mobility.

Due to the infectious mode of respiratory person-to-person transmission, location and population mobility continue to play a key role in the spread of COVID-19.^10^ Population mobility patterns inform risk of infectious disease exposure as well as highlight the varying and local non-pharmaceutical prevention mandates implemented during the pandemic. These mandates included limited business hours, reduced public location capacity, and stay-at-home orders. The time populations spend at locations where people are likely to interact—such as restaurants, health provider offices, grocery stores, and religious institutions or places of worship—are a dominant factor that has shaped this pandemic.^11-14^ These factors require inclusion in a model of disease spread.

The purpose of this study was to identify optimal spatial allocation methods for COVID-19 vaccines with the objective of minimizing reported COVID-19 cases. Utilizing GPS data from smart devices, we incorporated mobility and location factors into a mixed-effect Poisson model predicting the spread of COVID-19. When placed in an optimization framework, the resulting model is a convex math program, readily solvable through widely available software. By showing how to overcome major obstacles to better vaccine allocation, the methods we propose are both timely and practical.

## Methods

### Sample

This study utilized an econometric, repeated measure design with the 115 counties comprising Missouri as subjects, each with 26 weekly observations from January 2021 to July 2021. This study period was chosen to coincide with recommendation and release of vaccines to adult state residents.^8^ Data used for the study were collected from three primary sources. While this study was geographically limited to the state of Missouri, the location was decidedly generalizable due to (1) having both rural and urban locations and (2) the presence of COVID-19 prevention mandates in some, but not all counties.

### Measures

To build and assess vaccine allocation scenarios, the most appropriate outcome variable for this study was new weekly reported cases of COVID-19 per county, collected from the Missouri Department of Health and Senior Services.^15^ Weekly observations, rather than daily case counts, limited day-of-the-week reporting bias and more readily included retroactive data corrections.

Weekly vaccine uptake, for all available manufactured vaccines, among county residents was used as the study’s primary predictor and was collected from publicly available Missouri data.^16^ COVID-19 vaccine uptake was divided by two to reflect the two-dose vaccine requirement needed to reach recommended immunological protection.^15^

Aggregated and anonymized GPS data were collected from the data management firm Safegraph, LLC. This mobility data consisted of a rotating sample of 5-6% of the U.S. population who have consented to share data detailing time and location of visits outside the home.^17^ The data were stratified according to county of residence and then temporally across types of locations visited. Locations were organized by the North American Industrial Classification System. Approximately 250GB of uncompressed data were extracted from Safegraph, LLC prior to cleaning, organizing, and aggregating on the county level. We leveraged prior research to identify locations where risk of COVID-19 exposure would likely increase. These locations included restaurants/bars, health provider offices, grocery stores, education facilities, senior living facilities, retail locations, and religious institutions.^11-14^ Additional details regarding mobility data collection have been published elsewhere.^18,19^

Number of COVID-19 related deaths and hospital costs related to COVID-19 were estimated for use in analysis. These estimates were calculated by using the number of new COVID-19 cases along with the average national COVID-19 case-fatality-rate, average national rate of hospitalizations due to COVID-19, and hospital treatment costs of COVID-19 at time of respective observations.^1,20^

### Statistical Analysis

Statistical analysis of the data was completed in three phases. First, descriptive statistics identified temporal trends and variability among counties. Second, a mixed-effect generalized linear regression characterized the temporal correlation between COVID-19 vaccine distribution and COVID-19 case counts. The number of new COVID-19 cases was fit with a Poisson distribution to accommodate the non-negative count nature of the model outcome. A random effect was added to account for the nested nature of observations within the 115 counties. Fixed effects included average time spent at grocery stores, restaurants and bars, retail stores, healthcare delivery and service locations, education facilities, and senior living facilities per week per resident to reflect the variation in risk inherent among these locations. We also included the estimated total population of each county and the average distance traveled when residents visited locations outside the home.

In the last phase of analysis, the regression model was combined with a prescriptive optimization model for vaccine allocation. Given a limited supply of vaccines arriving across weeks 1, 2, …, *T* = 26, we allocated doses such that the total number of infections across counties 1, 2, …, *K* = 115 was minimized. During each week *t*, we chose 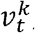, a number between zero and one representing a proportion of the population in county *k*. Letting *N*_*k*_ be the population size at county *k*, we allocated 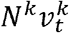 vaccines to county *k* during week *t*. We let *d*_*t*_ be the number of doses scheduled to arrive at the beginning of week *t* and assumed leftover inventory of vaccines could be carried from one week to the next.

The optimization utilized the Poisson model of disease spread to forecast case prevalence. We tied decision variables to the regression equation via the function 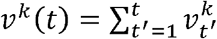, the proportion of the population at location *k* vaccinated from the beginning of the study period through week *t*. The covariates and their effects became data in the optimization. Let log*E* [*Y*_*tk*_ |*x*_*tk*_]=*a*_*tk*_ + β*ν*^*k*^*(t)* be the Poisson regression equation predicting the log of the expected number of cases *Y*_*tk*_ during week *t* in county *k* given covariate vector *x*_*tk*_, where β is the vaccination effect and *a*_*tk*_ represents the fixed and random effects associated with the remaining covariates. The expected number of cases was obtained by exponentiation.

The vaccine allocation problem was modeled as the following math program:

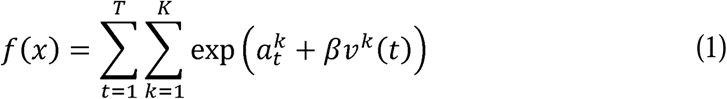

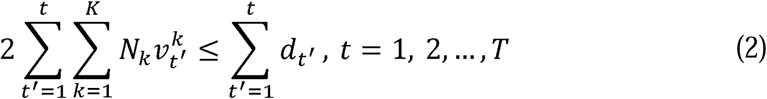

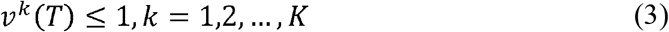

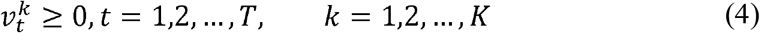

Objective function (1) was the expected total number of infections across all weeks and locations. Constraints (2) limited vaccine allocation in week *t* to the vaccines available in that week, which consisted of new deliveries plus any vaccines not allocated during previous weeks. Constraints (2) also required two vaccinations per individual. Constraints (3) limited the vaccines allocated to each county to be less than or equal to the county’s population. Constraints (4) required non-negative allocations. In practice, when 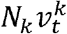 vaccines is not a whole number, then rounding down to the nearest integer resulted in a feasible allocation.

Because the math program is convex, any locally optimal allocation will also be a globally optimal allocation, meaning a better allocation does not exist.^9^ Consequently, any procedure guaranteed to identify locally optimal solutions in continuous-constrained optimization was adequate to find a globally optimal solution. We solved all instances of problem (1)-(4) with the Knitro solver.^21^ Proof of convexity is provided in the supplemental material.

To depict the effects of optimization, we devised nine scenarios by varying proportions of actual vaccine supply and resident mobility (time and distance traveled). Each scenario held supply and mobility factors at their actual levels, reduced them by 50%, or increased them by 100%. Each scenario represented an instance of the vaccine allocation optimization problem in equations (1)-(4). These scenarios provided different sets of mobility data to the objective and different supply schedules to the constraints, but held the fixed and random effects constant. In addition to identifying the minimum number of expected new COVID-19 cases in each scenario, we connected these figures to the expected number of COVID-19 related deaths and to expected hospital costs.

## Results

During the study period, a total of 173,656 COVID-19 cases were reported among Missouri counties for an average of 58.7 cases per week per county (SD 220.2). At the end of the study, counties had an average vaccination rate of 26.8% (SD =7.3%). Residents spent an average of 113.4 minutes (SD 64.2) when visiting senior living facilities, 99.4 minutes (SD 54.3) at healthcare facilities, 87.3 minutes (SD 26.2) at educational facilities, 43.1 minutes (SD 63.1) at grocery and food stores, 38.5 minutes (SD 16.1) at retail locations, and 37.2 minutes (SD 17.4) at restaurants and bars. Overall, residents traveled an average of 21.8 km (SD 13.3) to reach these locations during the study period.

Differences in county population sizes are depicted in the quantile map in Figure 1A. The quantile maps in Figures 1B and 1C show differences in average time at the specified locations and average distance traveled, respectively. Figure 1D displays the number of vaccines distributed among all counties at each week of the study, peaking at 354,894 during week 14.

**Figure 1.**
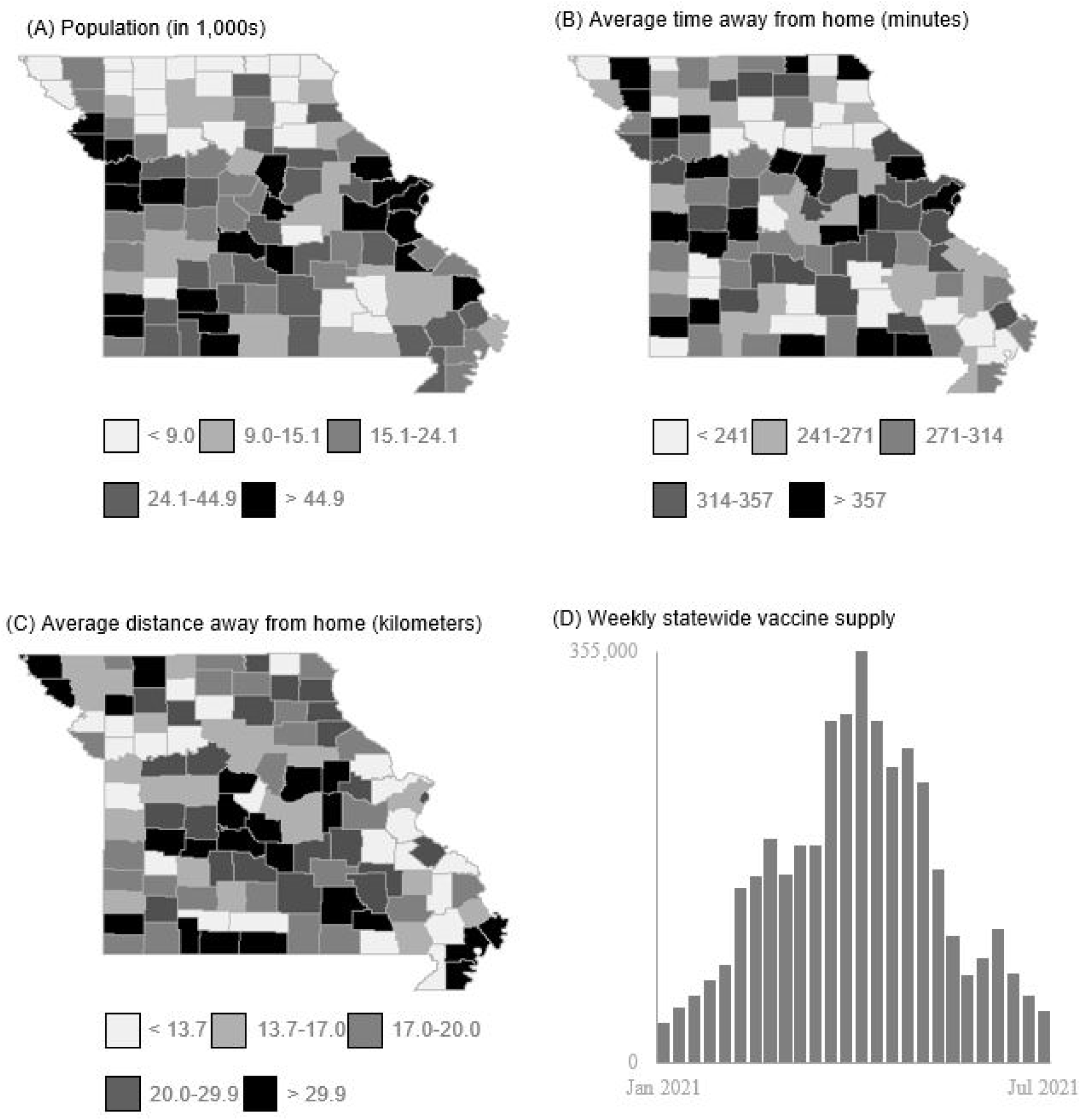
Descriptive quantile maps of (A) Missouri county populations (as of 2019), (B) average visit time (min) spent at a location outside the home (Jan 2021 – July 2021), (C) average distance (km) residents traveled to visit a location outside the home (Jan 2021 – July 20201), (D) weekly number of COVID-19 vaccines distributed among Missouri counties (Jan 2021 – July 2021)

Results of the mixed-effect regression, predicting number of COVID-19 cases, are detailed in Table 1 with the estimated variable coefficients expressed in log-link response. Each variable included in the model was shown to be significantly associated with the response variable. Relationships were as we expected: as the cumulative percent of vaccinated individuals increased, the number of new COVID-19 cases decreased; case prevalence increased with time spent at various commercial locations; and a county’s population and average distance away from home contributed to higher case counts.

**Table 1.** Fixed effect estimates predicting weekly COVID-19 cases among Missouri Counties from

|  | Coefficient estimate<sup>†</sup> | 95% Confidence interval | *p*-value |
| --- | --- | --- | --- |
| % Vaccinated | -2.488 | -2.552, -2.423 | <0.001 |
| Time spent at grocery/food stores (min) | 0.001 | 0.000, 0.001 | <0.001 |
| Time spent at restaurants/bars (min) | 0.014 | 0.013, 0.015 | <0.001 |
| Time spent at retail locations (min) | 0.016 | 0.015, 0.016 | <0.001 |
| Time spent at healthcare locations (min) | 0.004 | 0.003, 0.004 | <0.001 |
| Time spent at education locations (min) | 0.001 | 0.011, 0.012 | <0.001 |
| Time spent at senior living facility (min) | 0.004 | 0.004, 0.004 | <0.001 |
| Distance traveled from residence (km) | 0.371 | 0.356, 0.385 | <0.001 |
| Population<sup>‡</sup> | 0.912 | 0.709, 1.114 | <0.001 |

<sup>†</sup> log-link response, <sup>‡</sup> scaled around minimum and maximum values

For each of the nine mobility-supply scenarios, Figure 2 compares the performance of optimal allocation against a population-based allocation, where vaccines were distributed only according to population size. In the 100% mobility and 100% supply scenario (Fig. 2, Scenario 5), the state of Missouri’s actual allocation policy was used as a second benchmark. In this scenario, we predict spatial optimization of vaccine allocation would have averted 72,781 COVID-19 cases, averted 1,201 COVID-19 related deaths, and saved $54,893,389 in COVID-19 related hospital costs. The optimal vaccine allocation was nine percentage points more effective, based on averted cases, than the population-based allocation and eight percentage points more effective than Missouri’s actual allocation. The largest disparity between optimized allocation and population-based allocation was seen when resident mobility was doubled and vaccine supply was halved (Fig. 2, Scenario 7). Under these parameters, optimized allocation averted twice as many cases as the population-based allocation method. Even under the most favorable parameters, with mobility halved and vaccine supply doubled (Fig. 2, Scenario 3), the number of cases averted by the optimal allocation was six percentage points higher than the number averted by the population-based allocation.

**Figure 2.**
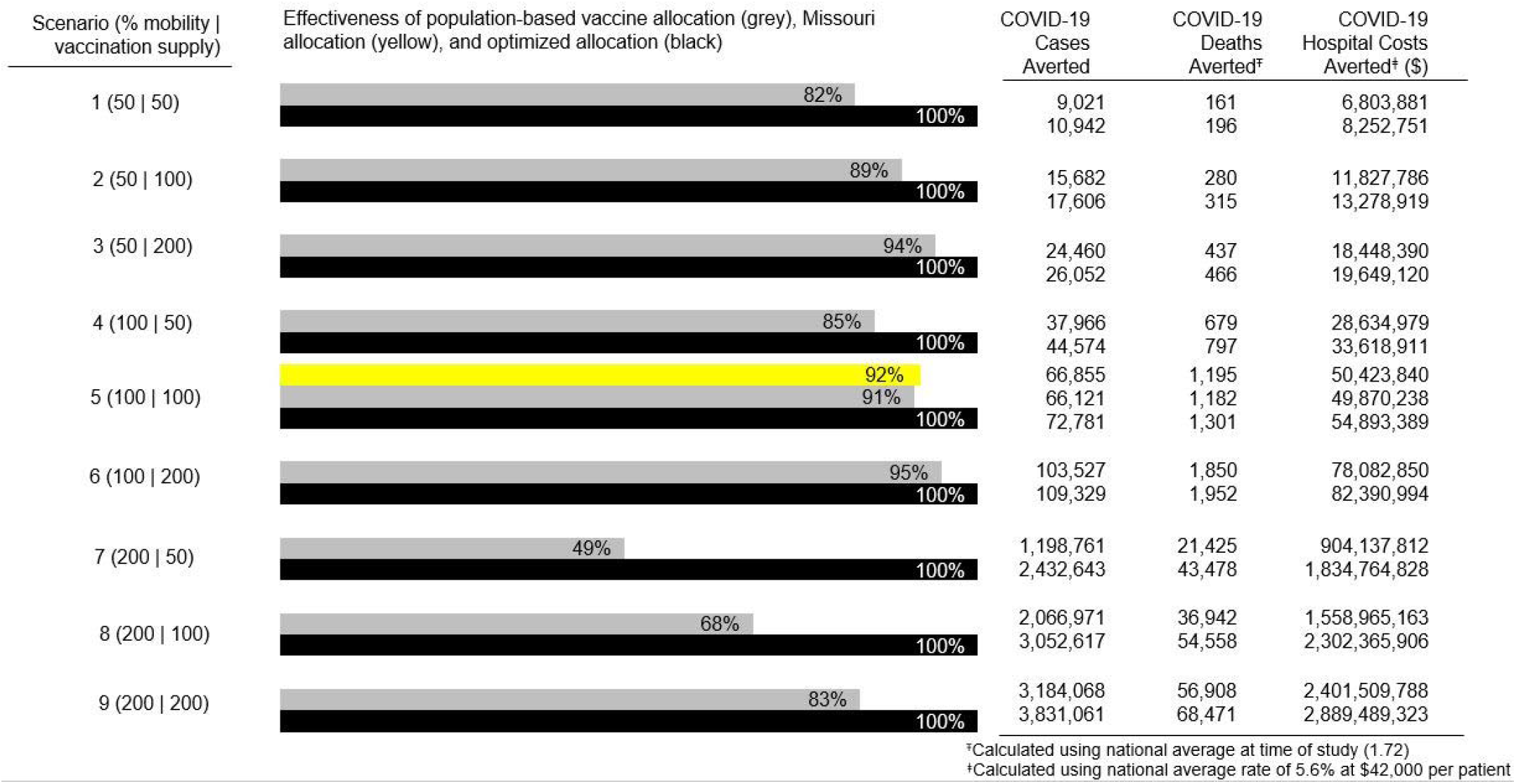
Effectiveness of population-based allocation and Missouri state allocation compared to spatially optimized allocation under nine different scenarios of varying geographic mobility and vaccine supply.

Finally, we examined the value of vaccines across time in an optimal allocation policy. For each of the nine scenarios, Figure 3 displays the dual variables associated with weekly supply constraints (3). Due to large differences in dual values, we display the figures in three charts with identical horizontal scales but with different vertical scales for each mobility level. A value in Figure 3 can roughly be interpreted as the decrease in case count that would have resulted from one additional vaccine available for allocation during a particular week. This information is a unique byproduct of mathematical optimization and cannot be obtained through other means.

**Figure 3.**
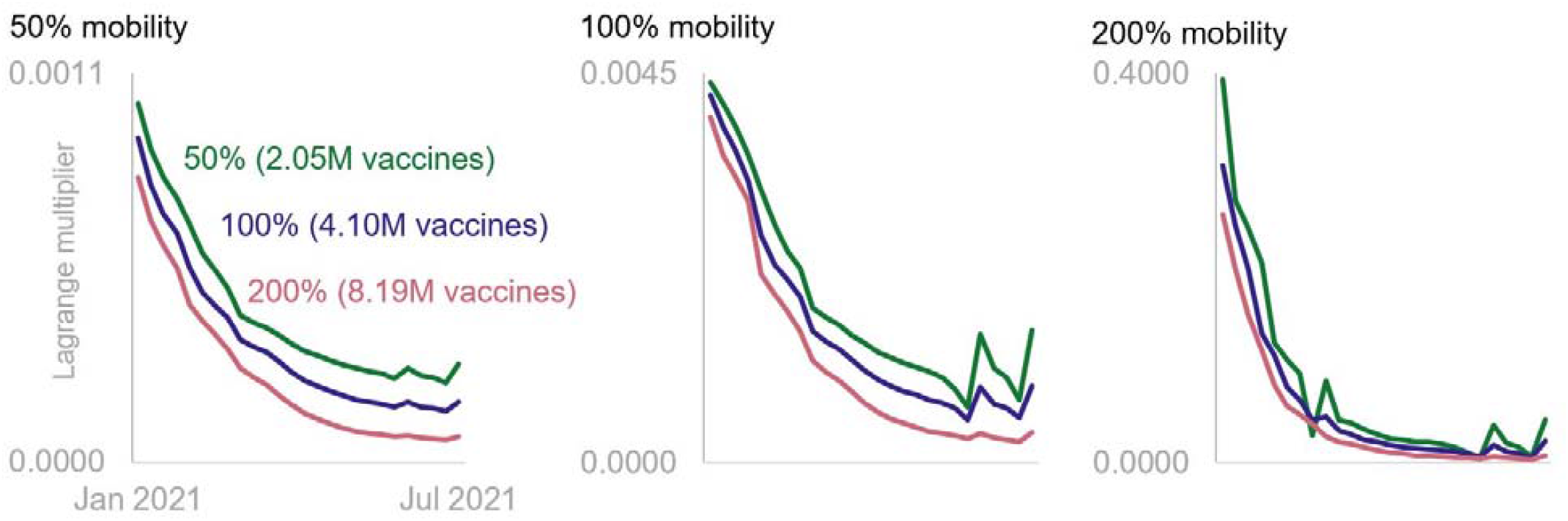
Changes in temporal value of COVID-19 vaccines from January 2021 to July 2021 under differing geographic mobility

## Conclusions

The purpose of this study was to understand the impact of spatially optimal COVID-19 vaccine allocation in Missouri. Results suggest optimal allocation would have markedly improved health outcomes, reducing the number of cases by 8%. Beyond this overall finding, we report two other important results.

First, across all scenarios in Figure 3, vaccines were generally more valuable when they were allocated earlier rather than later. For example, when mobility was 50% and supply was 200% (Fig. 2, Scenario 3), an additional vaccine had more than 12 times the impact in early January 2021 than it would have had toward the end of June 2021. This difference increased to more than 73 times when mobility was 200% and supply was 50% (Fig. 2, Scenario 7). Because COVID-19 infections grew at an exponential rate, a given number of vaccines was more effective at slowing disease spread in the early parts of a pandemic than the same amount would have been later. The kinks in each series are related to variations in the supply schedule. When the number of vaccines available for allocation during a particular week was lower than the supply the week before and the week after, additional vaccines during that week were more valuable.

Second, across all time periods, the value of a vaccine increased substantially as mobility of the population increased. For instance, when supply was at 50%, the dual variable corresponding to the week-one supply constraint increased by more than a factor of four when mobility moved from 50% to 100%, then by an additional 90 times when mobility increased to 200 percent. That is more than a 38,000% increase from low to high mobility. This enormous difference points to the importance of mobility in curbing a pandemic. It is not that a vaccine’s ability to inoculate somehow increases as individuals spend more time outside of their residences and venture further away from their homes. Rather, as Figure 2 shows, the number of infections to be averted is orders of magnitude higher, and thus the potential for a vaccine to decrease disease spread is also much higher.

Our work provides an important public health tool for the future. In the face of new COVID-19 variants, our analysis can be used to guide the distribution of limited supplies of resources. Further, as we prepare for the possibility of other pandemics, this research lays a foundation for the integration of important environmental factors into predictive disease models and prescriptive optimization tools.

Though the literature on COVID-19 vaccine allocation is young, the same realism-tractability challenge faced by many fields is present here. Optimization models that integrate location, mobility, and disease dynamics are better representations of reality than those that do not, but they are significantly more difficult to solve.^22,23^ Our work considered the roles of mobility and location in disease progression and also yielded provably optimal vaccine allocation policies. We demonstrated how optimal policies could have averted infections, deaths, and hospital costs in the U.S. state of Missouri during the first half of 2021, a period when vaccine supplies were low, and COVID-19 infections continued to increase. Across a range of scenarios, we showed the potential for an optimal allocation of vaccines to improve upon policies based on population size. We found that the benefits of optimal allocation increased dramatically in scenarios with higher mobility and fewer vaccines. But even when mobility was low and supplies were more abundant, optimal allocation of vaccines still led to reductions in case prevalence, fatalities, and hospital costs.

To conceptualize findings and propel future research, several study limitations were identified. Due to data availability, this study worked under the assumption that distribution of vaccines equated to administration of vaccines. However, news sources revealed that at times vaccines go unused.^24,25^ In addition to including geographic mobility, it may be beneficial for future studies to consider collective community beliefs and attitudes surrounding likelihood of vaccine uptake. While this study also gives an estimate of COVID-19 deaths and hospitalization costs, these values are based on national averages and, like infection rates, are likely a product of geographic variation.^26,27^ Additional studies would benefit from deeper examination of these variables and the role they play in optimal vaccine allocation policies.

## Supporting information

optimization equation description

## Data Availability

Portions of data used for this study are publically available through the Missouri Department of Health and Senior Services COVID-19 Dashboard. Mobility data contained within this study was collected and shared with researchers by Safegraph LLC through a member-approved academic research consortium.

## Acknowledgments

This work was partially supported by the Sinquefield Center for Applied Economic Research at Saint Louis University.

