## Supplementary material for "Spatial Optimization to Improve COVID-19 Vaccine Allocation": optimization equation description

To show that problem (1)-(4) is a convex program, the objective function must be a convex function and the constraints must form a convex set. As the constraints are all linear, they trivially form a convex set. We demonstrate that objective function $f$ is convex by showing that the Hessian matrix, which contains all second partial derivatives of $f$, is positive definite. We begin with first partial derivatives. The partial derivative of $f$ with respect to $v_{t}^{k}$ is

$$\frac{\partial f}{\partial v_{t}^{k}}= \sum_{\tilde{t}=t}^{T} \beta exp\left. \left( a_{\tilde{t}}^{k}+ \beta v^{k}\left( \tilde{t} \right) \right. \right)$$

Further differentiating with respect to $v_{\bar{t}}^{\bar{k}}$ gives

$$\frac{\partial^{2}f}{\partial v_{t}^{k}\partial v_{\bar{t}}^{\bar{k}}}=\left\{ \begin{aligned} \sum_{\tilde{t}=\max\left\{ \tilde{t}, t \right\}}^{T} \beta^{2} exp\left. \left( a_{\tilde{t}}^{k}+ \beta v^{k}\left( \tilde{t} \right) \right. \right), \text{ }k=\bar{k,} \\ 0, \text{otherwise.} \end{aligned} \right.$$

Note that the second partial derivative is strictly positive if $k=\bar{k}$, otherwise it is zero. When $k=\bar{k}$, the summation goes from the larger of $t$ and $\bar{t}$ up to $T$. Consequently, the Hessian matrix can be constructed as a block diagonal matrix, where each block has special structure. For a given $k$, define the block

$$P_{k}=\left[ \begin{matrix} \frac{\partial^{2}f}{\partial v_{1}^{k}\partial v_{1}^{k}} & \frac{\partial^{2}f}{\partial v_{1}^{k}\partial v_{2}^{k}} & \cdots& \frac{\partial^{2}f}{\partial v_{1}^{k}\partial v_{T}^{k}} \\ \frac{\partial^{2}f}{\partial v_{2}^{k}\partial v_{1}^{k}} & \frac{\partial^{2}f}{\partial v_{2}^{k}\partial v_{2}^{k}} & \cdots& \frac{\partial^{2}f}{\partial v_{2}^{k}\partial v_{T}^{k}} \\ \vdots& \vdots& \ddots& \vdots\\ \frac{\partial^{2}f}{\partial v_{T}^{k}\partial v_{1}^{k}} & \frac{\partial^{2}f}{\partial v_{T}^{k}\partial v_{2}^{k}} & \cdots& \frac{\partial^{2}f}{\partial v_{T}^{k}\partial v_{T}^{k}} \end{matrix} \right]\text{ .}$$

In the $t^{\text{th}}$ row, the first $t$ elements are identical. Then, denoting by $\boldsymbol{0}$ the $T\text{-by-}T$ matrix of zeros, the Hessian is

$$H\left( x \right)=\left[ \begin{matrix} P_{1} & \boldsymbol{0} & \cdots& \boldsymbol{0} \\ \boldsymbol{0} & P_{2} & \cdots& \boldsymbol{0} \\ \vdots& \vdots& \ddots& \vdots\\ \boldsymbol{0} & \boldsymbol{0} & \cdots& P_{K} \end{matrix} \right]\text{ .}$$

Objective function $f$ is convex if $H$ is positive definite. One way to demonstrate that $H$ is positive definite is to show that its pivots are all positive. To do this, use Gaussian elimination to put $H$ in echelon form by performing the following row operations on each block: for each row $t$ except the last row, replace row $t$ with row $t$ less row $t+1$. In the new row, if $\bar{t}>t$, all terms cancel and the result is zero. Otherwise, the result is strictly positive. It follows that each block is a lower triangular matrix of strictly positive values, and thus the pivots, which constitute the diagonal, are also positive. Thus, the block is positive definite. Performing identical operations on all blocks demonstrates that $H$ is positive definite, and thus $f$ is convex.
